# Change in outbreak epicenter and its impact on the importation risks of COVID-19 progression: a modelling study

**DOI:** 10.1101/2020.03.17.20036681

**Authors:** Oyelola A Adegboye, Adeshina I Adekunle, Anton Pak, Ezra Gayawan, Denis HY Leung, Diana P Rojas, Faiz Elfaki, Emma S McBryde, Damon P Eisen

**Author notes:** Corresponding author: Oyelola A. Adegboye. Australian Institute of Tropical Health and Medicine, James Cook University, 1 James Cook Drive, Douglas, Queensland, Australia, 4814.

## Abstract

The outbreak of Severe Acute Respiratory Syndrome Coronavirus-2 (SARS-CoV-2) that originated in the city of Wuhan, China has now spread to every inhabitable continent, but now theattention has shifted from China to other epicenters, especially Italy. This study explored the influence of spatial proximities and travel patterns from Italy on the further spread of SARS-CoV-2 around the globe. We showed that as the epicenter changes, the dynamics of SARS-CoV-2 spread change to reflect spatial proximities.

---

As at March 10th 2020, 109 countries and territories have confirmed at least one case of COVID-19 and a total number of 113,702 COVID-19 cases have been confirmed globally – 80,924 in China and 32,778 outside China [1]. Human population movement plays an important role in the spread of infectious diseases. Although China has served as the epicenter of the current SARS-CoV-2 outbreak, real-time phylogenetic analysis [2] suggests that the virus was introduced to most African countries and to Latin America from people traveling from Italy. Migration has remained the major source of concern for the current COVID-19 outbreak and most countries have focused on China as the likely source of any importation. The movement of people between China and sub-Saharan Africa has increased rapidly over the last decade [3, 4], the spread of COVID-19 to the African continent has been more related to the current COVID-19 outbreak in Italy. There has been much attention on the importation of infectious diseases such as Ebola, Tuberculosis, Malaria or viral hepatitis from Africa to Europe [5, 6]. Conversely, there are few reported cases of importing diseases from the European countries **to** Africa. This, perhaps, makes most African countries to focus their COVID-19 surveillance efforts on travelers from China without much attention paid to the possible importation from other countries.

In addition to the phylogenetic analysis, a number of other circumstantial factors support the introduction of the virus to other countries, as it appears more likely that the introduction of COVID-19 to South America occurred from Italy rather than from China. First, since January 23, 2020, China has increased their containment measures and decreased the number of outbound international flights. Second, early febrile airport screening mostly targeted travelers coming from Asia overlooking those from other parts of the world – including travelers from Europe; an approach that turned out to have limited effectiveness [7]. Third, as most of the infections by SARS-CoV-2 are asymptomatic and mild [8-10], it is more likely that infected carriers could enter a country without being detected by the temperature screening at the airports. Lastly, Italian Brazilians are the largest group of foreigners with full or partial Italian ancestry outside Italy, with São Paulo being the most populous city with Italian ancestry in the world. In this paper, we investigated how the growing epidemic in Italy affects the spread of the epidemic around the world by using measurements of spatial proximity and travel volume between countries.

## Methods

This study is based on the number of confirmed cases of COVID-19 in each country until March 10^th^, 2020 obtained from World Health Organisation (WHO) situation reports [1] and real-time online COVID-19 monitoring sites [11, 12]. The air travel data between countries were obtained from the Official Airline Guide (OAG) database (www.oag.com). Preliminary analysis includes Pearson’s correlation analysis to examine the association between the number of COVID-19 cases and travel influx (and arrival time) from the source country. Spatial data reveal the degree of dependency among observations in geographical space [13, 14]; consequently, spatial dependence measures such semi-variogram and Local Indicator of Spatial Association (LISA) [15] were used to detect spatial patterns in COVID-19 data. For the spatial weights, we used Queen-style contiguity 1st order nearest neighbour (i.e., two countries are neighbouring if they share common borders or a point). Additionally, we developed a stochastic meta-population model of global spread using the 2018 IATA flight data for travel volumes between each country. The country-specific compartmental model is an adaptation of the classical SEIR (Susceptible-Exposed-Infectious-Recovered) model based on reported dynamics of the novel coronavirus [16, 17] for global spread of COVID-19 (see the appendix for model details and www.pandemic.org.au for deterministic version) [18]. We used the cases on WHO situation report [1] to initialise the number of people infected in Italy and assuming same for China to determine global risk from these two epicenters under the same initial infection seed. Each country’s risk of importation was classified into quartiles: slight risk (<25%), moderate risk (25%-50%), high risk (50%-75%) and extreme risk (>75%).

## Results and Discussion

Up until the February 21^st^ Italian outbreak, all reported European cases were imported from China (Figure 1) [19]. The disease has now (as at March 10^th^) spread to 46 (excluding Vatican City and Liechtenstein) out of the 53 countries within the WHO European region [1]. Italy has 9,172 confirmed cases and 463 deaths as of March 10^th^. The region alone accounted for 46% of the total cases outside of China. There are currently three COVID-19 epicenters –China, Italy, and Iran, which are mostly responsible for introducing the virus globally (Figure 1).

**Figure 1:**
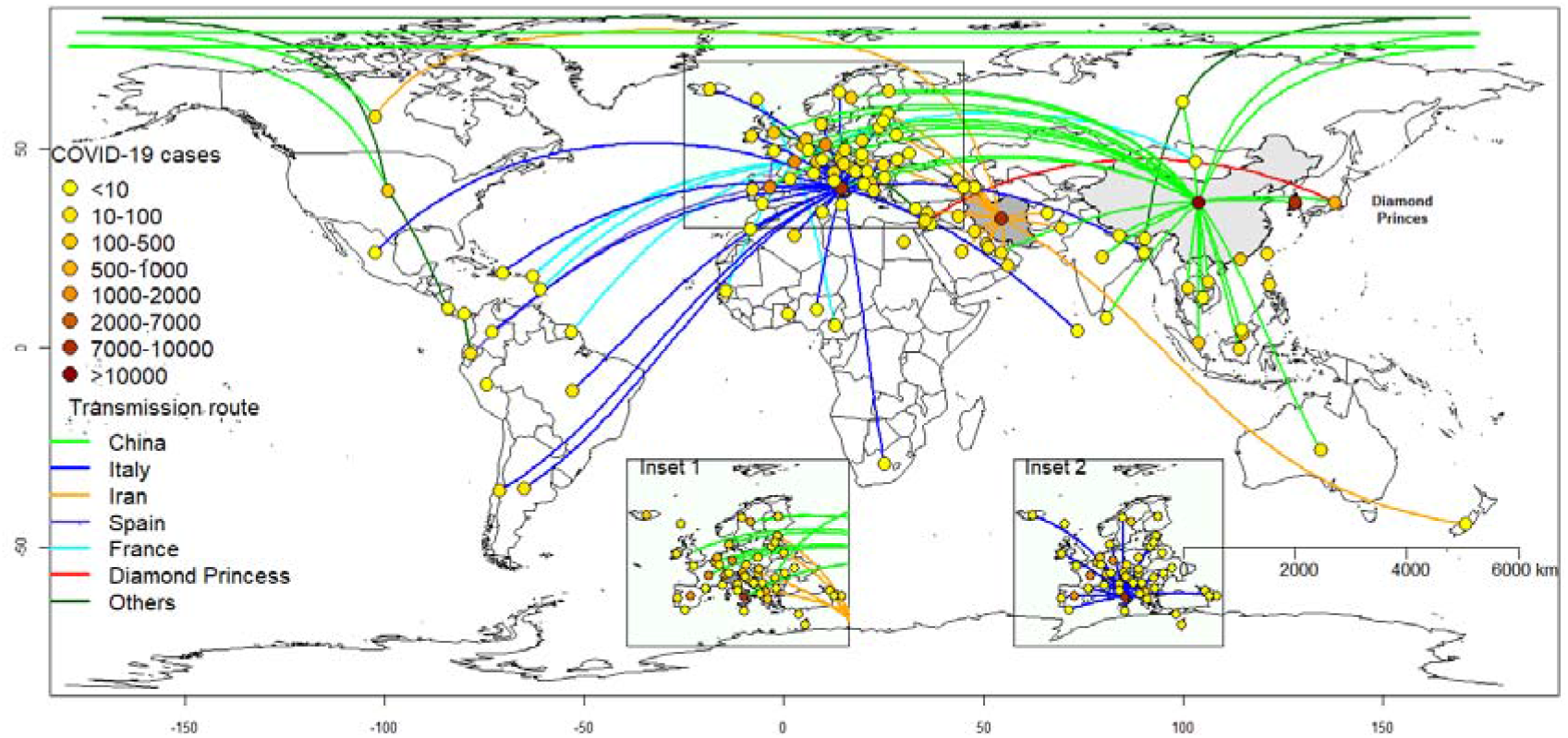
Transmission routes of COVID-19 as at March 7^th^ 2020. The lines represent transmission routes from the source of COVID-19 into a country. Inset 1: European cases originating from China (green lines) and Iran (orange lines). Inset 2: Cases originating from Italy.

We present in Figure 2, the distribution of time elapsed since 21^st^ February when the first imported case of COVID-19 from Italy was reported in each country against the geographical distance from Italy. The outbreak in Italy is highly sporadic in the last three weeks. While the shortest arrival time (after the Italy outbreak) was 5 days to Austria, Switzerland, Brazil and Algeria, the virus spread to 44 countries within 17 days (median, 11 days). The global arrival times for three epicenters from the first reported cases in Wuhan, China are shown in Figure S1.

**Figure 2:**
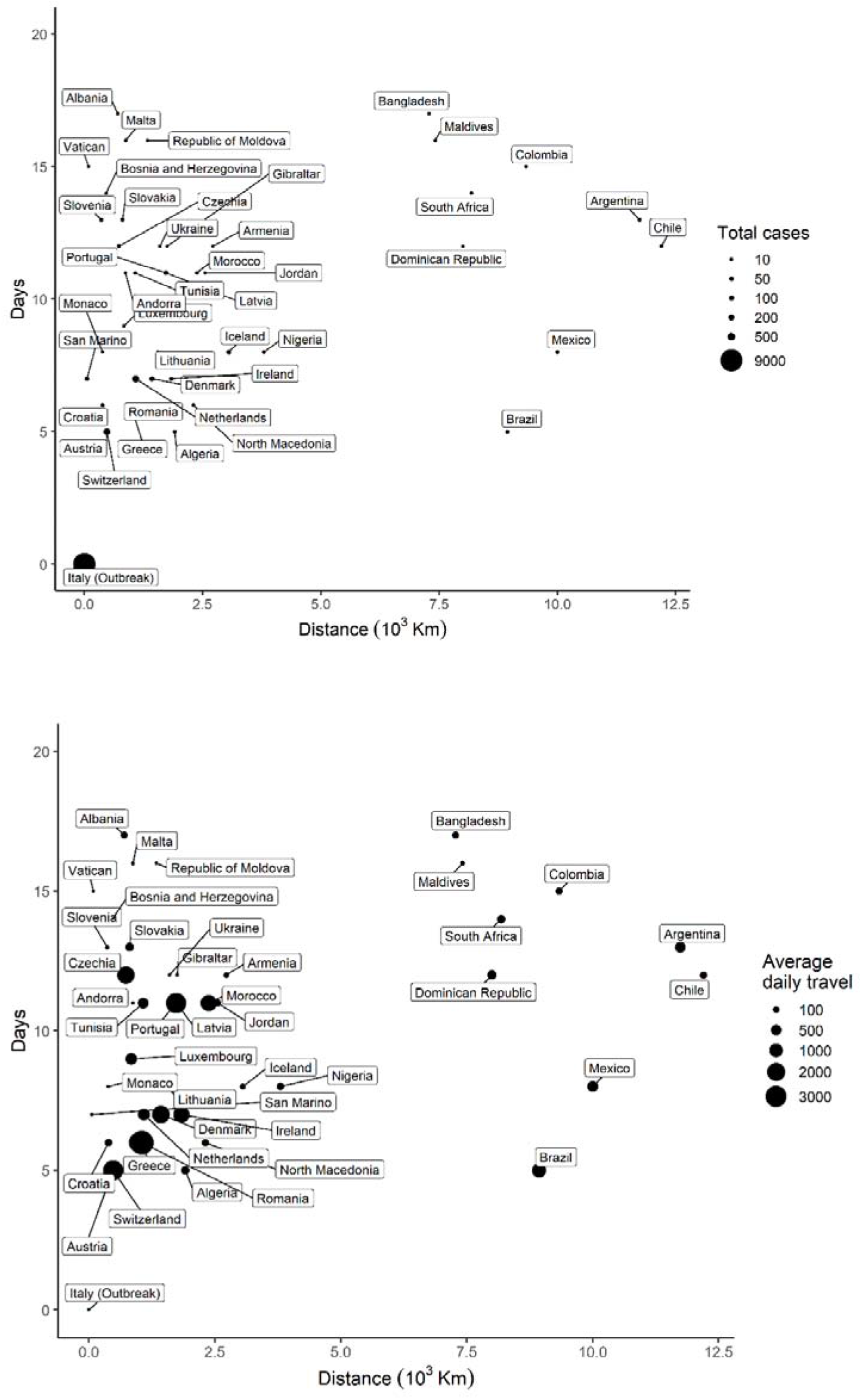
Distribution of arrival times of COVID-19 cases against geographic distance from the source (Italy). The dots are proportional to the size of, (A) total cases, and (B) travel influx from the source country.

The Pearson’s correlation analysis (r= -0.43, p=0.004) suggest negative association between disease arrival time and number of cases for cases imported from Italy. That is, countries with shorter introduction time have more number of cases. There was significant association between the number of cases and daily travel influx from Italy (r=0.39, p=0.011). The apparent spatial autocorrelation in the COVID cases within Europe (Moran’s Index=0.310, p=<0.001) is also significant (Figure S2). Notably, France and Italy are located in the right upper quadrant, while Switzerland, Austria and Slovakia are on the left upper quadrant, indicating a positive and negative spatial autocorrelation pattern respectively. Additionally, semi-variograms indicated spatial autocorrelation of the disease incidence exists up to a distance of 120 decimal degrees for Italy (Figure S3). Using bivariate LISA analysis, we found evidence of spatial interaction between COVID-19 cases and arrival time as well as travel influx (Moran’s I=0.340. This suggests that spatial variations within European region were non-random, exhibiting effects of neighbouring interactions and travel influx.

We simulated COVID-19 spread comparing two outbreak epicenters – Italy and China. Figure 3 shows the relative risk of importation from the two epicenters. The results show that whereas the Asia-Pacific is more at risk (high risk to extreme) from China epicenter, Europe, South-America and Africa are more at risk from the Italian epicenter. High population density and highly interconnected transportation networks connecting tourism hubs in Northern Italy with major European cities have made it extremely difficult to contain and reduce the number of infections. This aligns well with our prediction of extreme risks of case importations from Italy to other European, while Latin American and African countries have high risks (Figure 3). In the case of Italy, our modelling results could provide an argument for the Italian government instituting a national quarantine and travel restrictions on March 9^th^, 2020 to mitigate consequent spread of infection. Based on the evidence form the Chinese policies of containment and quarantine that showed considerable effectiveness [20], these interventions should slow the spread of the disease within Europe and abroad.

**Figure 3:**
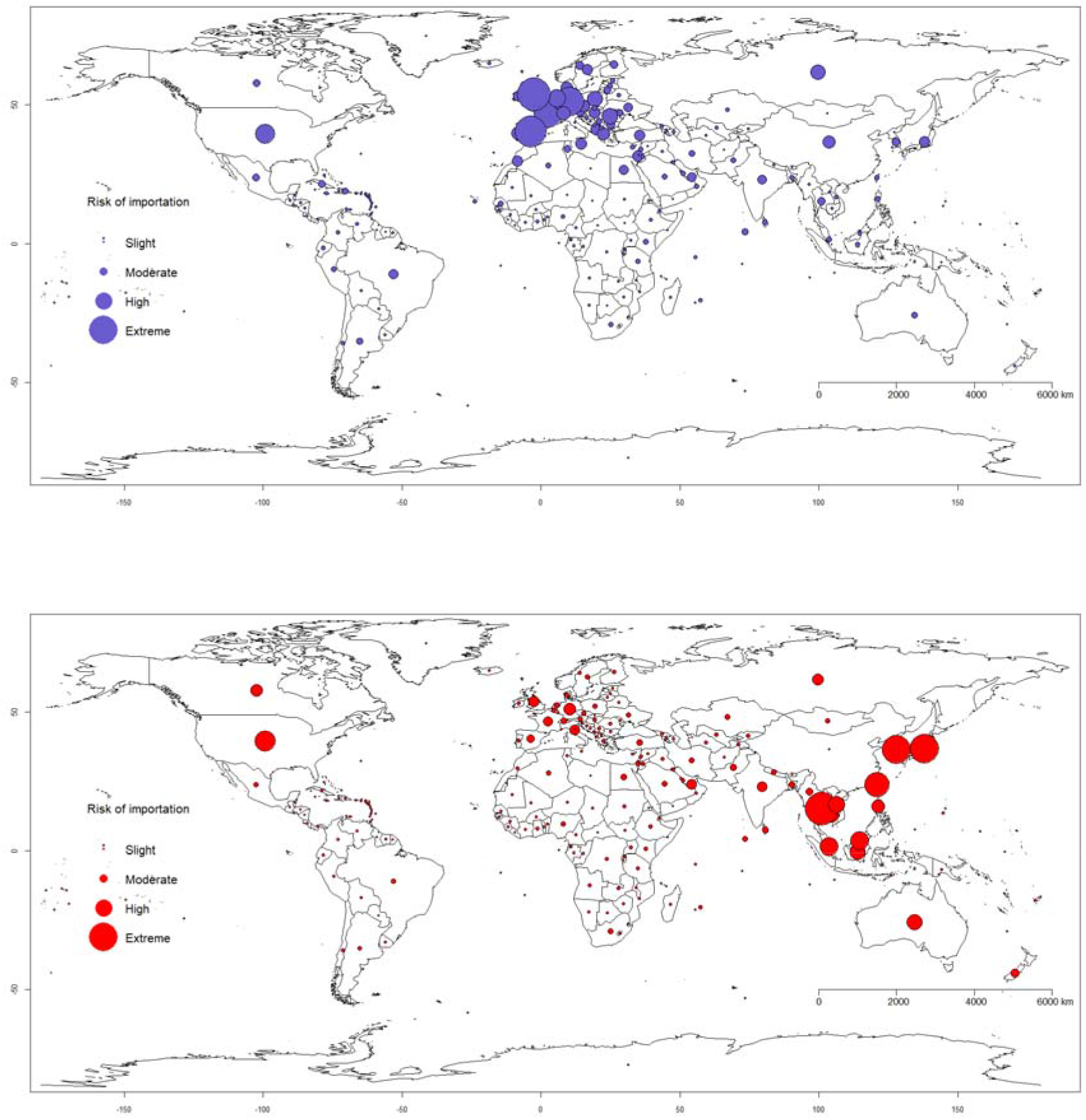
Global risk of importation of COVID-19 from, (A) Italy (purple dot) and (B) China (red dot) based on air travel influx from source country. The dots are proportional to the cumulative relative risk that an infected individual will be arriving at a specific country for an epicenter. Risk of importation: slight risk (<25%), moderate risk (25%-50%), high risk (50%-75%) and extreme risk (>75%).

Although most African countries have focused their COVID-19 surveillance efforts on travelers from China, our analysis shows that this approach may need to be reconsidered as the number of cases in Europe soar, making the risk of importation to Africa higher from European countries than from China through stronger transportation connectivity and migration flows (especially, with North African countries). As at now, Italy has put itself under lockdown [21] and other countries such as Australia and the USA have restricted flights from Italy [22, 23]. As new epicenters are emerging, countries are required to adapt quickly and adjust their containment measures to reduce the spread of infection.

## Conclusion

In summary, we estimated the risk of importation from two major epicenters to other countries and showed that spatial proximity and mobility are important factors that fuel disease importation. This analysis illustrates the potential development of the infection spread and may be useful in countries’ epidemic preparedness including large-scale interventions such as travel restrictions and quarantine.

## Data Availability

The data used in this should is publicly available on WHO website

https://www.who.int/emergencies/diseases/novel-coronavirus-2019/situation-reports

## Appendix 1

**Table S1:** State variable and parameter descriptions

| Variable | Description |
| --- | --- |
| $S$ | Susceptible class |
| $E_1$ | Latent stage not infectious |
| $E_2$ | Latent stage infectious |
| $I_1$ | First stage of symptomatic |
| $I_2$ | Second stage of symptomatic |
| $R$ | Recovered class |

| Parameter | Description | Values (references) |
| --- | --- | --- |
| $\beta_1, \beta_2, \beta_3$ | Transmission rates for $E_2, I_1, I_2$ classes | 0.3125, 0.5, 0.176 ([17]) |
| $\sigma_1$ | First stage incubation rate | 0.3125 ([17]) |
| $\sigma_2$ | Second stage incubation rate | 0.5 ([17]) |
| $\gamma_1$ | First stage of recovery | 0.5 ([17]) |
| $\gamma_2$ | second stage of recovery | 0.176 ([17]) |
| $c_h$ | COVID-19 case fatality | 0.018 ([24]) |
| $\mu$ | Death rate | Not used |

**Figure S1.**
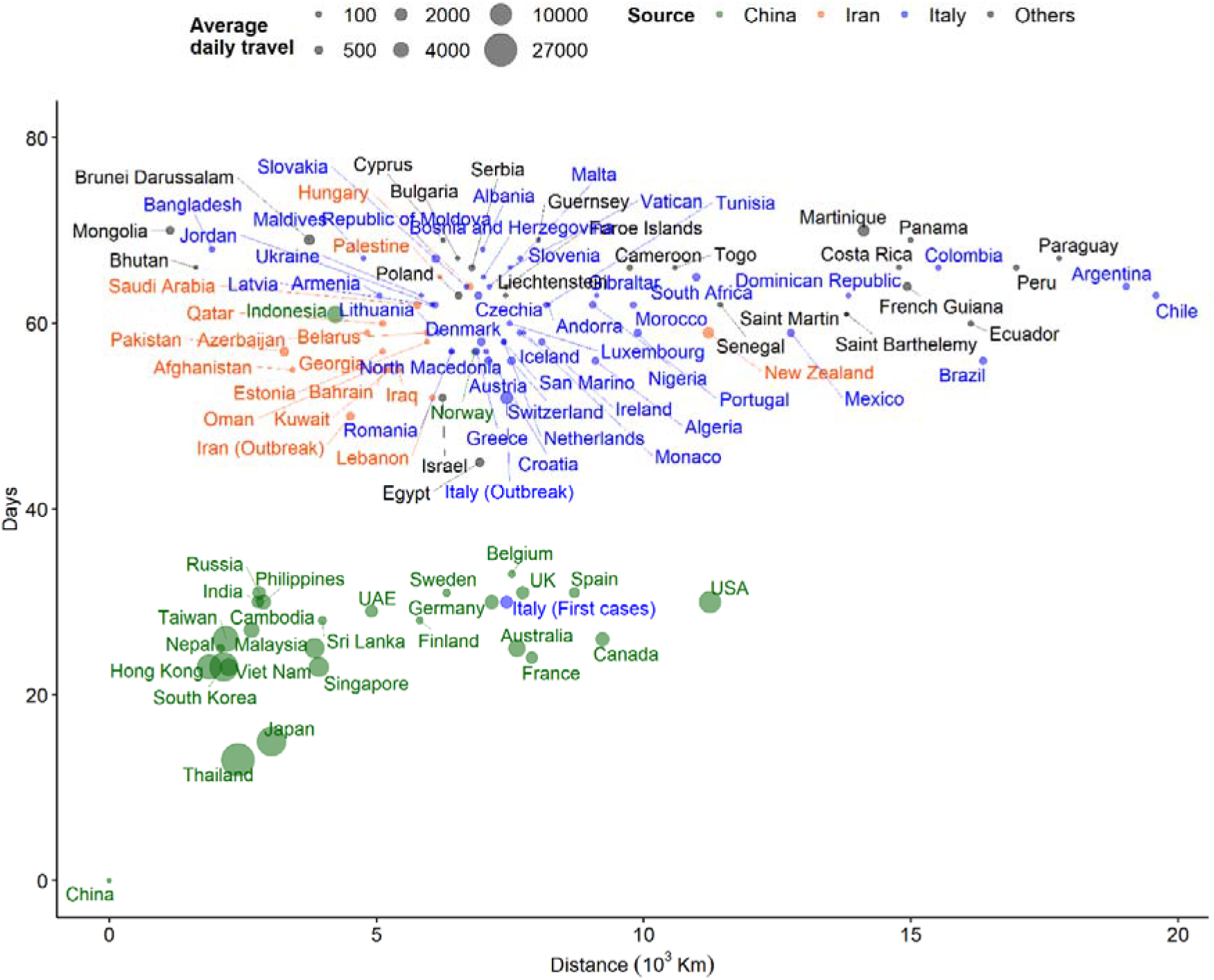
Distribution of COVID-19 arrival times (days from first reported cases in China) and distance from China. The size of the dot is proportional to the daily air travellers between China and other countries as at March 10^th,^ 2020.

**Figure S2.**
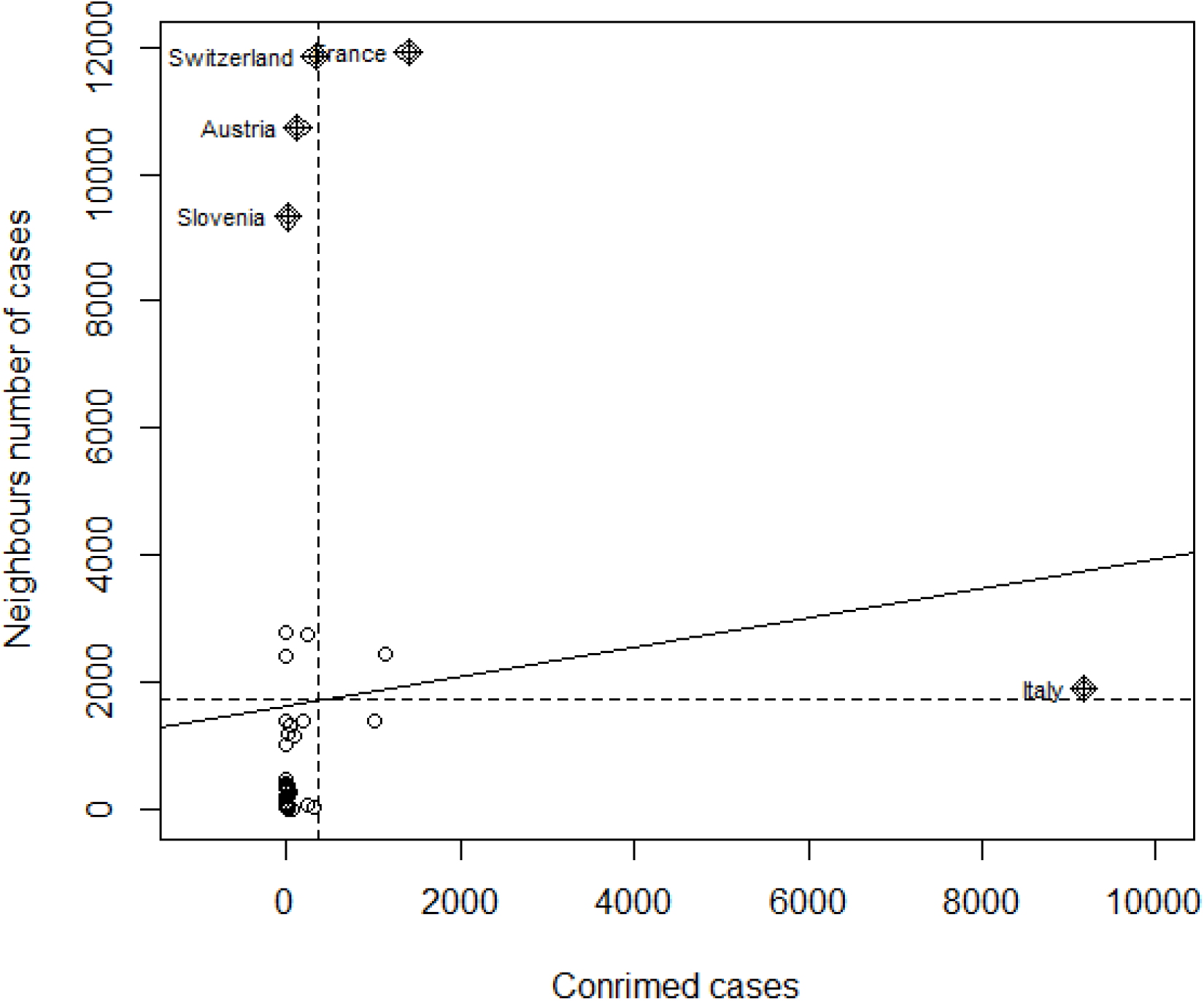
Moran’s Index for COVID-19 cases in Europe

**Figure S3:**
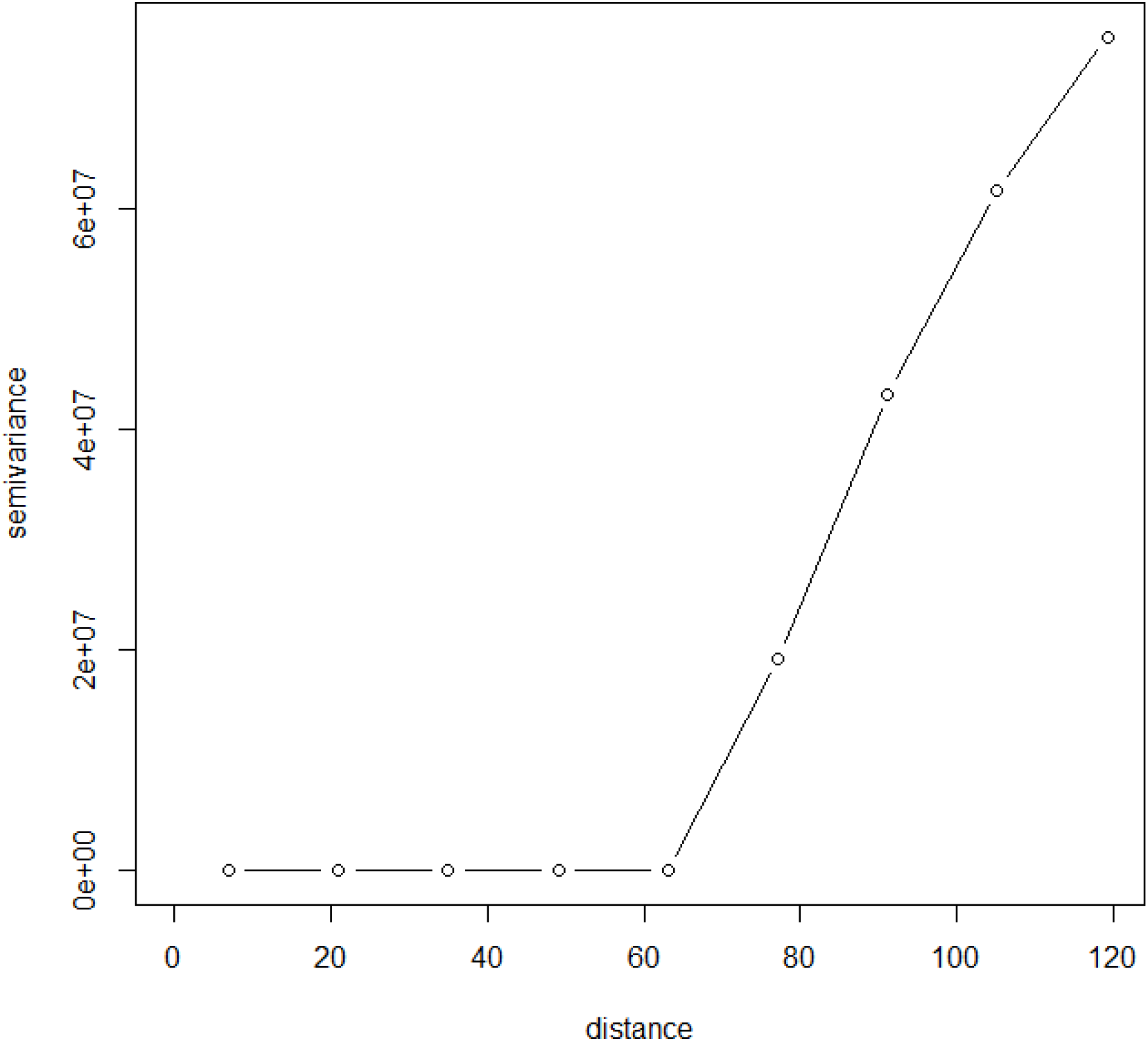
Semi-variogram showing spatial autocorrelation for COVID-19 data up 0.000 decimal degrees (∼1.2km) for Italy and …for China.

**Figure S4:**
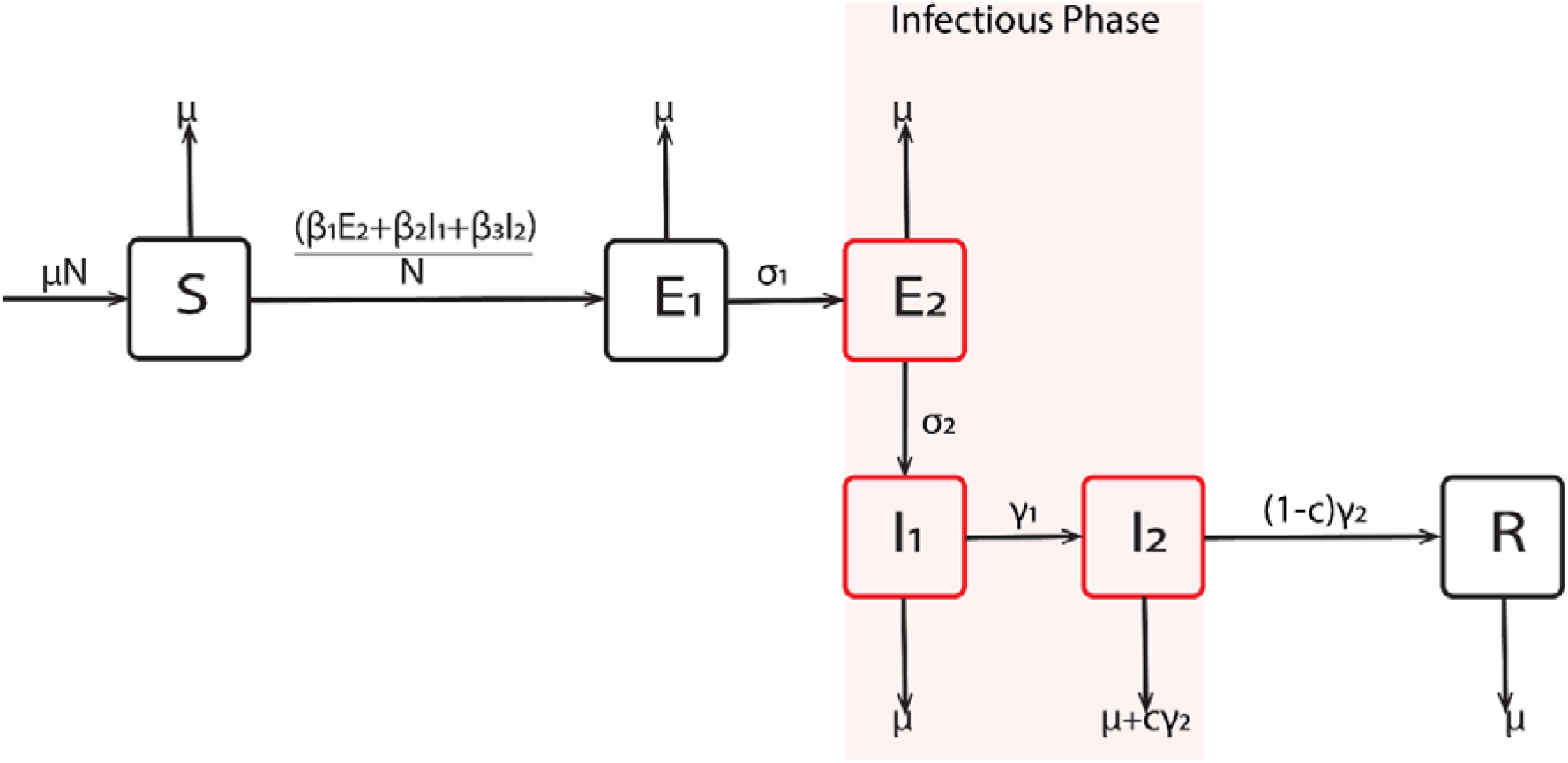
Schematic diagram of the transmission dynamics

## Appendix 2: Mathematical Model details

The infectious classes are late latency *E*_2_ and the two stages of symptomatic infectiousness (*I*_1_ and *I*_2_). Patients either recover and moved to R class or die and are replaced to ensure constant population. The two-country dynamical model is as shown below:

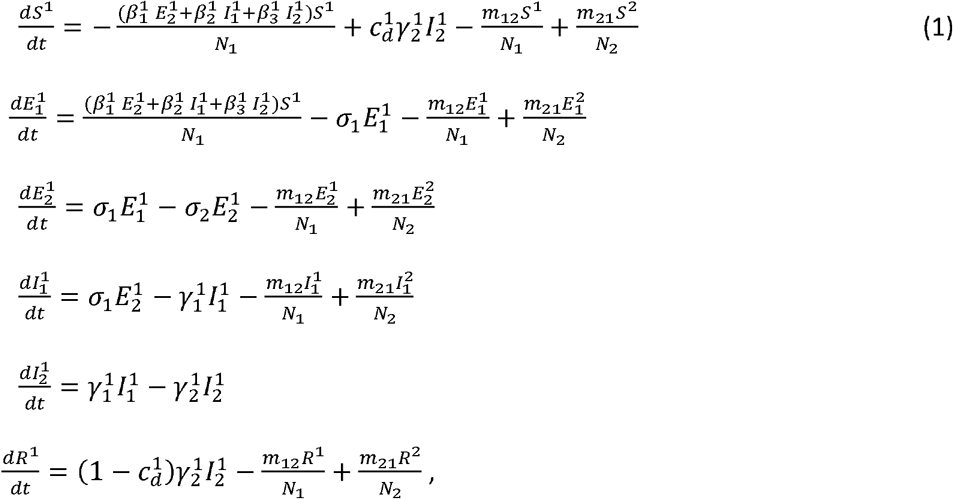

where superscript indicates country.

This model is extended to all countries and coded in R [25] using the infectious disease node of the Australia Nectar Research cloud (www.nectar.org.au) as individual-based model with a binomial distribution of the number of people experience an event at a specific time step. The events are infection, migration, emigration, recovery and death due to COVID-19. We neglected natural death, as this does not affect our result. We further assumed that no countries except the epicenters are experiencing outbreak. Hence the basic reproduction number at the epicentres is *R*_0_ = 2.68. Each country COVID-19 dynamics follows the schematic representation in Fig S4.

